# Chronic COVID-19 Syndrome and Chronic Fatigue Syndrome (ME/CFS) following the first pandemic wave in Germany – a first analysis of a prospective observational study

**DOI:** 10.1101/2021.02.06.21249256

**Authors:** C Kedor, H Freitag, L Meyer-Arndt, K Wittke, T Zoller, F Steinbeis, M Haffke, G Rudolf, B Heidecker, HD Volk, C Skurk, F Paul, J Bellmann-Strobl, C Scheibenbogen

**Affiliations:** Institute of Medical Immunology, 13353 Berlin, Charité - Universitätsmedizin Berlin, corporate member of Freie Universität Berlin, Humboldt Universität zu Berlin and Berlin Institute of Health, Germany; Experimental and Clinical Research Center, Max Delbrueck Center for Molecular Medicine and Charité - Universitätsmedizin Berlin; Department of Infectious Diseases and Respiratory Medicine, Charité - Universitätsmedizin Berlin; Department of Cardiology, Charité - Universitätsmedizin Berlin; Berlin-Brandenburg Center for Regenerative Therapies (BCRT), Charité University Medicine Berlin, Germany

**Keywords:** COVID-19, Chronic COVID-19 Syndrome, Chronic Fatigue Syndrome, Myalgic Encephalomyelitis, SARS-CoV-2, long-COVID-19

## Abstract

**Objective:** Characterization of the clinical features of patients with persistent symptoms after mild to moderate COVID-19 infection and exploration of factors associated with the development of Chronic COVID-19 Syndrome (CCS).

**Methods:** Setting: Charité Fatigue Center with clinical immunologists and rheumatologist, neurologists and cardiologists at Charité University hospital.

Participants: 42 patients who presented with persistent moderate to severe fatigue six months following a mostly mild SARS-CoV-2 infection at the Charité Fatigue Center from July to November 2020.

Main outcome measures: The primary outcomes were clinical and paraclinical data and meeting diagnostic criteria for Chronic Fatigue Syndrome (ME/CFS). Relevant neurological and cardiopulmonary morbidity was excluded.

**Results:** The median age was 36.5, range 22–62, 29 patients were female and 13 male. At six months post acute COVID-19 all patients had fatigue (Chalder Fatigue Score median 25 of 33, range 14–32), the most frequent other symptoms were post exertional malaise (n=41), cognitive symptoms (n=40), headache (n=38), and muscle pain (n=35). Most patients were moderately to severely impaired in daily live with a median Bell disability score of 50 (range 15–90) of 100 (healthy) and Short Form 36 (SF-36) physical function score of 63 (range 15-80) of 100. 19 of 42 patients fulfilled the 2003 Canadian Consensus Criteria for myalgic encephalomyelitis/chronic fatigue syndrome (ME/CFS). These patients reported more fatigue in the Chalder Fatigue Score (p=0.006), more stress intolerance (p=0.042) and more frequent and longer post exertional malaise (PEM) (p=0.003), and hypersensitivity to noise (p=0.029), light (p=0.0143) and temperature (p=0.024) compared to patients not meeting ME/CFS criteria. Handgrip force was diminished in most patients compared to healthy control values, and lower in CCS/CFS compared to non-CFS CCS (Fmax1 p=0.085, Fmax2, p=0.050, Fmean1 p=0.043, Fmean2 p=0.034, mean of 10 repeat handgrips, 29 female patients). Mannose-binding lectin (MBL) deficiency was observed frequently (22% of all patients) and elevated IL-8 levels were found in 43% of patients.

**Conclusions:** Chronic COVID-19 Syndrome at months 6 is a multisymptomatic frequently debilitating disease fulfilling diagnostic criteria of ME/CFS in about half of the patients in our study. Research in mechanisms and clinical trials are urgently needed.

## INTRODUCTION

Infection with severe acute respiratory syndrome coronavirus type 2 (SARS-CoV-2) poses a major threat for developing chronic morbidity. While older patients or those with risk factors have a high risk of severe or critical COVID-19 (Corona Virus Disease) mortality and morbidity, in about 80% of cases COVID-19 is mild according to WHO criteria. Soon there were reports, however, of patients with persistent symptoms following mild COVID-19 referred to as long COVID.^1 2^ Frequent symptoms that were reported are fatigue, impaired physical and cognitive function, headache, breathlessness, palpitations and many other symptoms, impairing activities of daily living in many patients.^3-7^ A recent publication of a patient survey of long COVID in younger patients described diverse symptoms with fatigue, post-exertional malaise (PEM), and cognitive dysfunction as most frequent requiring a reduced work schedule in almost half and inability to work in 22% of patients.^3^ PEM describes an intolerance to mental and physical exertion, which triggers an aggravation of symptoms typically lasting for more than 14 hours up to several days^8^.

Long-term health consequences following mild COVID-19 are largely unknown yet but have been feared based on observations from SARS-CoV-1. Here many patients were reported who developed a severe post-infectious syndrome with persistent fatigue, muscle pain, shortness of breath and mental symptoms independent of illness severity.^9^ Various pathogens including Epstein Barr Virus (EBV), enteroviruses, and dengue viruses are known to trigger chronic fatigue syndrome/myalgic encephalomyelitis (ME/CFS) in a subset of patients.^10^ It is unclear yet, if pathomechanisms of post-infectious fatigue syndromes may be different depending on the pathogen.

ME/CFS is a debilitating chronic disease with a worldwide prevalence of 0.3 to 0.8%.^11^ Profound mental and physical fatigue, sleep disturbance, and chronic pain are key symptoms of ME/CFS. The best discriminating symptoms distinguishing ME/CFS from chronic fatigue in multiple sclerosis were flu-like symptoms and the intolerance to mental and physical exertion triggering PEM for more than 14 hours.^12^ ME/CFS is classified by the World Health Organization (WHO) as neurological disease with G93.3 in the International Classification of Diseases (ICD). Although the pathomechanism is not well characterized yet, there is ample evidence of immune, autonomous nervous system and metabolic dysregulation.^13^ There is emerging evidence that post-infectious ME/CFS has an autoimmune mechanism and dysfunctional autoantibodies to natural regulatory antibodies against adrenergic receptors were described.^13-15^

We report here on the first results of our ongoing study initiated at Charité in July 2020 to characterize patients with persistent fatigue and other symptoms following mild to moderate COVID-19 and to assess if they meet diagnostic criteria for ME/CFS. Due to the complexity of symptoms patients were comprehensively evaluated by a team from various disciplines including clinical immunology, rheumatology, neurology, cardiology, and pneumology with long experience in diagnosing ME/CFS (https://cfc.charite.de). Our findings confirm initial concerns that COVID-19 leads to persistent fatigue syndromes in a subset of younger individuals following mild to moderate infection. We describe here the clinical characteristics of the Chronic COVID-19 Syndrome (CCS) at month six following acute infection with a subset of patients fulfilling diagnostic criteria of ME/CFS.

## MATERIAL AND METHODS

### Study protocol

The primary objective of this monocentric prospective observational study is to characterize patients contacting the Charité Fatigue Center with persistent fatigue after COVID-19 prospectively and determine if they fulfill diagnostic criteria for ME/CFS. Patients were selected based on a screening questionnaire, specifying COVID-19 diagnosis and symptoms including mild to moderate COVID-19 according to WHO criteria and persistent symptoms. Inclusion criteria were symptoms of moderate to severe fatigue and exertion intolerance, neurocognitive impairment, and pain six months post infection in the absence of relevant respiratory, neurological or psychiatric comorbidity. A subset of patients was already seen at months three and four, and reevaluated at month six. COVID-19 had been diagnosed by PCR (polymerase chain reaction) or serology (SARS-CoV 2 IgG/IgA) or if PCR was not performed and serology was negative clinically due to typical initial symptoms including loss of smell and taste (n=3). All patients signed informed consent before study assessment. This study is part of the Pa-COVID-19 study of Charité and approved by the Ethics Committee of Charité Universitätsmedizin Berlin in accordance with the 1964 Declaration of Helsinki and its later amendments (EA2/066/20). Between July, 16 and November, 27 a total of 57 patients presented at our outpatient clinics. Patients were excluded from this study in the presence of relevant comorbidities or preexisting fatigue, absence of confirmed COVID-19 or evidence of organ dysfunction. Further patients were not included in this report in case of pending cardiopulmonary or neurological assessment. Thus, we report here on the results of the cross-sectional analysis at month six in a total of 42 patients.

### Diagnostic criteria and symptom assessment

Severity of mental and physical fatigue was assessed using Chalder Fatigue Score. Disability and daily physical function was assessed by Bell disability scale and Short Form Health Survey (SF-36 Version 1). The Bell disability scale is scored from 0 (very severe, bedridden constantly)–100 (healthy).^16^ Frequency and the severity of PEM symptoms were assessed according to Cotler *et al*.^8^. Symptoms of autonomic dysfunction were assessed by the Composite Autonomic Symptom Score (COMPASS 31).^17^ Depression and sleepiness were assessed by Patient Health Questionnaire 9 (PHQ9) and Epworth Sleepiness Scale (ESS). According to PHQ9 patients were classified as minimal depressive symptoms (1-4), mild depressive symptoms (5-9), moderate depressive symptoms (10-14), moderately severe depressive symptoms (15-19), or severe depressive symptoms (20-27). According to ESS patients were classified as no evidence of sleep apnea 0–9, possible mild to moderate sleep apnea 11-15, >16 possible severe sleep apnea. One patient with a PHQ9 score of >20 who is in ongoing psychosomatic evaluation was excluded from this report.

A diagnosis of ME/CFS was based on Canadian Consensus criteria (CCC) and exclusion of other diseases, which may explain chronic fatigue and potential confounding comorbidities.^18^ In contrast to the original classification and in accordance with the studies of L. Jason and colleagues a minimum of 14 hours of PEM instead of 24 hours was required for diagnosis of ME/CFS.^8^ Key symptoms of CCC were quantified using a 1–10 scale. All data was recorded using a REDCap database.

### Functional studies, imaging and laboratory values

The handgrip strength was assessed using an electric dynamometer assessing maximal and mean force of ten times repeated maximal pulls (Fmax1 and Fmean1) and a second assessment 60 minutes later (Fmax2 and Fmean2, ref Jäkel B *et al*. in revision, 2021). Blood pressure and heart rate was assessed sitting, and after two, five and ten minutes standing. Postural tachycardia syndrome (POTS) is defined as increase of >30 bpm or over 120 bpm over ten minutes standing.^19^ Most patients had already neurological, pulmonary and cardiac assessment before referral to our outpatient clinic without evidence for relevant impairment or comorbidity. In patients who reported moderate to severe difficulties with breathing, chest computer tomography (CT) and pulmonary function tests were performed. Patients who reported severe cognitive impairment or severe headache got a further comprehensive neurological assessment from our neurology team. Patients with sitting or postural tachycardia or elevated NT-proBNP got a cardiologic examination including ECG, 24h ECG and echocardiography from our cardiology team. Laboratory parameters including CBC, lymphoycte subsets, IL-8 in erythrocytes, mannose binding lectin (MBL), CrP, immunoglobulins, ANA, ENA, C3/4, anti-TPO, TSH, fT3/4, ferritin, creatinine, liver enzymes, ACE, NT-pro BNP were determined at the Charité diagnostics laboratory (Labor Berlin GmbH, Berlin, Germany).

### Patient and Public Involvement

The German facebook group of patients with long COVID contacted us first in July 2020 sharing their observations (https://c19langzeitbeschwerden.de/). The study design was developed based on frequency, type and severity of symptoms reported and discussed with the patient group. The possibility for local patients to participate in our study was communicated on their website.

### Statistical analysis

We performed the statistical analysis with Excel GraphPad Prism 6.0. We used non parametric Mann-Whitney test to analyze differences between groups. For comparison of pulse and blood pressure at different time points we used Wilcoxon test. To exclude sex-related differences in hand grip strength and blood pressure, here, we evaluated only female subjects. For correlation analysis we calculated non parametric Spearman correlation coefficients. A p-value of <0.05 was considered as statistically significant. Due to multiple testing p-values are considered descriptive without adjustments for multiple comparisons.

## RESULTS

### Patient characteristics and disability

We report here on 42 patients who presented to the Charité fatigue center with post COVID-19 fatigue at month six. **Table 1** summarizes demographic characteristics of the study population. Most patients had mild COVID-19 (n = 32) and ten had moderate COVID-19 due to pneumonia.

**Table 1:** Demographic and baseline clinical characteristics

|  | CCS/CFS (n=19) |  | CCS (n=23) |  |
| --- | --- | --- | --- | --- |
|  | median | range | median | range |
| age | 41 | (24-62) | 36 | (22-57) |
| sex (female/male) | 14 / 5 |  | 15 / 8 |  |
| BMI | 24,31 | (18.1-31.8) | 22.49 | (18.0-36.2) |
| Bell | 40 | (20-80) | 50 | (30-90) |
| Chalder | 26 | (20-31) | 23 | (14-32) |
| PHQ9 | 12 | (3-19) | 11 | (2-18) |
| ESS | 9 | (1-21) | 9 | (1-17) |

CCS/CFS = chronic COVID-19 syndrome/chronic fatigue syndrome; CCS = chronic COVID-19 syndrome without fulfilling CFS criteria; BMI = body mass index (kg/m<sup>2</sup>), PHQ9 = Patient Health Questionnaire 9; ESS = Epworth Sleepiness Scale.

None of the patients required oxygen or mechanical ventilation and all but 3 patients were ambulatory. **Supplementary Table S1** shows the ten most frequent initial symptoms of COVID-19 reported by patients.

19 patients fulfilled the Canadian Consensus Criteria (CCC) for ME/CFS. These patients were referred to as Chronic COVID-19 Syndrome/CFS (CCS/CFS), the other patients as CCS. Most CCS patients (18 of 23) did not fulfill ME/CFS criteria due to a duration of PEM less than 14 hours. Further eight of 23 patients did not fulfill the criterium neurological/cognitive symptoms. Based on PHQ9 we have no evidence of severe mental health issues in our study cohort. (**Table 1**). Two patients had an ESS score of >16, which may indicate sleep apnea. In one patient ESS score at month three was only three when she presented already with similar symptom severity, the other patient had an ESS score of 17. These patients were referred to further investigation at the sleep medicine ambulance.

The majority of patients were severely impaired in daily live with a median Bell disability score of 40 (range 20–80) of 100 (refers to healthy) and a SF-36 physical function of 65 (range 15-90) of 100 (refers to healthy) in CCS/CFS and a Bell score of 50 (range 30–90) and SF-36 physical function score of 60 (range 25–90) in CCS. CCS/CFS patients reported lower SF-36 scores for vitality, role limitations and social functioning (p = 0.060, 0.005, 0.082, respectively, **Fig. 1**). According to the Bell scale patients with a score of 30–50 are able to perform light work 2–5 hours a day (n=28), thus requiring a reduced work schedule or inability to work. Patients with a Bell score of 20 (n=3) are confined to bed most of the day.

**Fig. 1.**
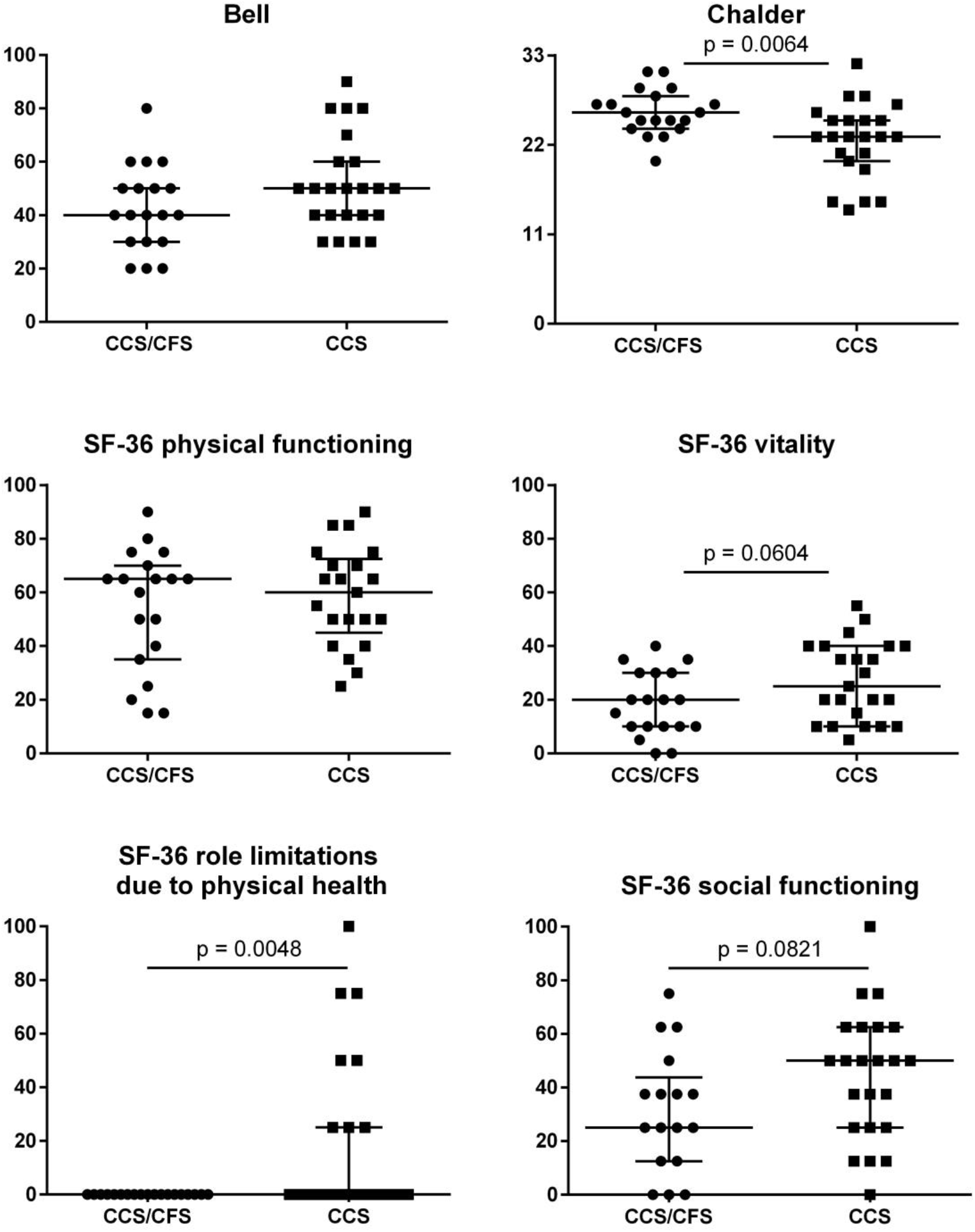
Severity of fatigue and disability. Bell disability score, Chalder fatigue score and SF-36 physical function, vitality, role limitations and social function in 19 CCS/CFS and 23 CCS patients. Median, range and single values are shown.

### Symptom severity

The leading symptom of fatigue was assessed by Chalder Fatigue Questionnaire (CFQ). Patients classified as CCS/CFS reported a significantly higher Chalder Fatigue Score with a median of 26 (range 20-31) compared to the other patients with a median 23 (range 14–32, p=0.006, **Fig. 1**). The severity of PEM as the cardinal symptom of ME/CFS was the strongest discriminatory symptom. CCS/CFS patients had significantly more frequent episodes of PEM (p=0.016). While according to diagnostic criteria all CCS/CFS patients had PEM of 14 hours or more, CCS patients reported PEM of less than 14 hours in 18 of 23 patients, most patients reported PEM lasting between 2–10 hours (n=15), only one patient had no PEM as shown in **Fig. 2**. Symptom severity was assessed on a scale of 1–10 (none to severest). **Table 2** shows the frequency and severity of all symptoms at month six in patients classified as CCS/CFS and CCS.

**Fig. 2.**
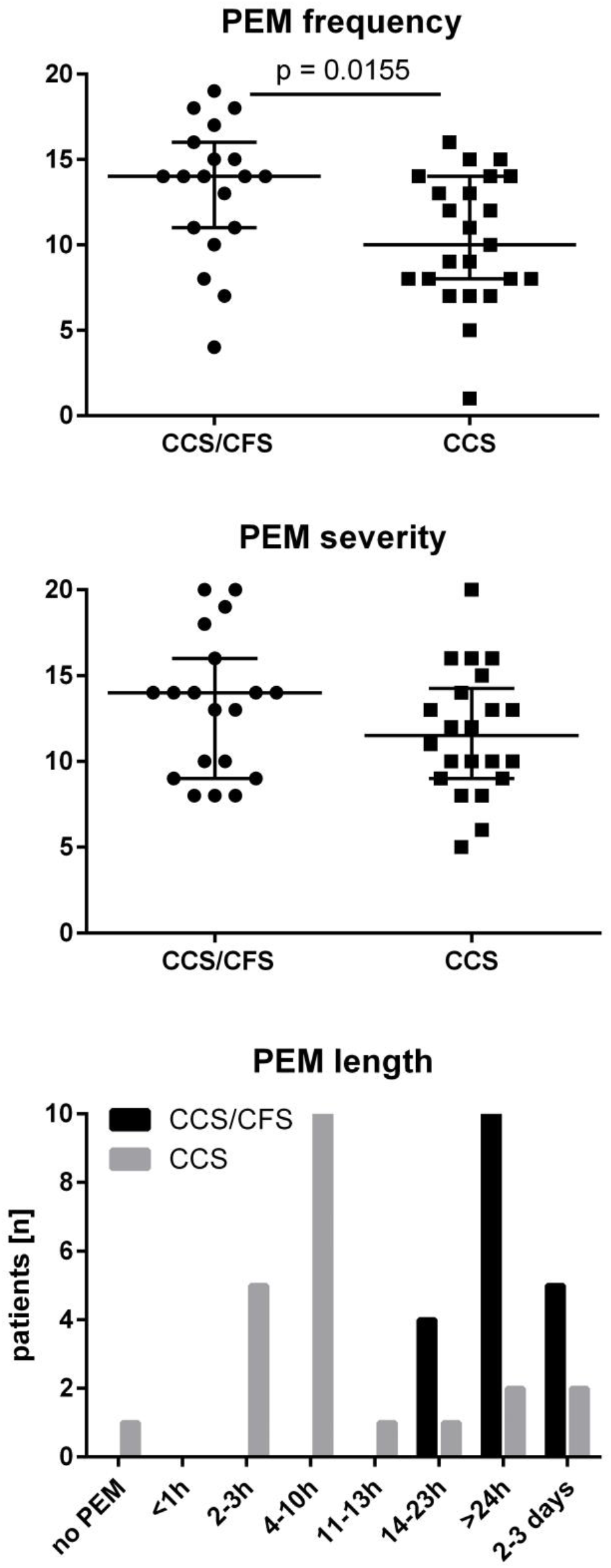
Frequency, severity and length of post exertional malaise (PEM) Frequency and severity of PEM was assessed on a five items scale with 0 – 20 points (none to severest) and the length in seven categories (from <1h to 2 – 3 days) according to Cotler.^8^ Median, range and single values are shown in 19 CCS/CFS and 23 CCS patients.

**Table 2:**
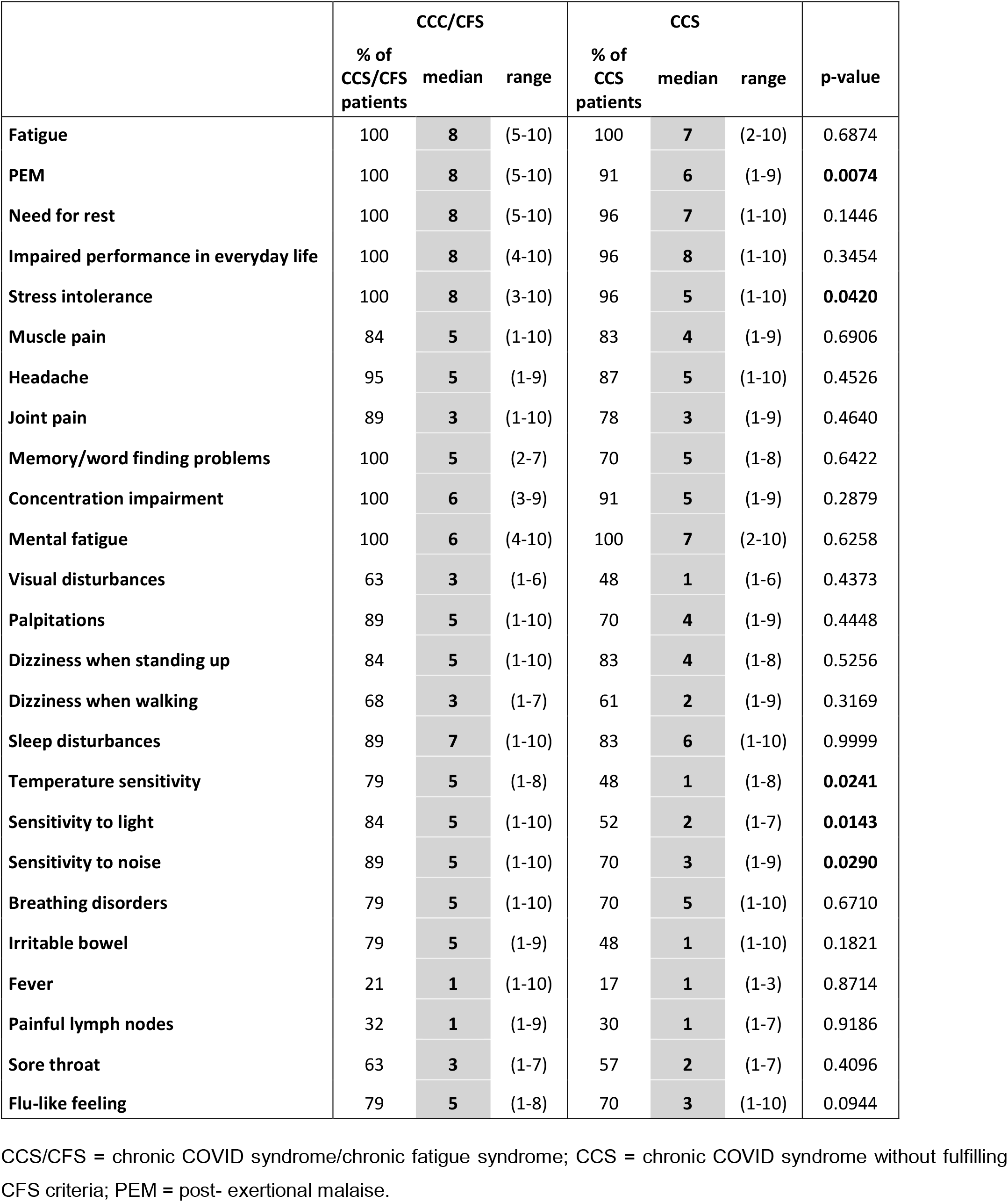
Frequency and severity of symptoms at month 6

|  | CCC/CFS |  |  | CCS |  |  | p-value |
| --- | --- | --- | --- | --- | --- | --- | --- |
|  | % of<br>CCS/CFS<br>patients | median | range | % of<br>CCS<br>patients | median | range |  |
| Fatigue | 100 | 8 | (5-10) | 100 | 7 | (2-10) | 0.6874 |
| PEM | 100 | 8 | (5-10) | 91 | 6 | (1-9) | **0.0074** |
| Need for rest | 100 | 8 | (5-10) | 96 | 7 | (1-10) | 0.1446 |
| Impaired performance in everyday life | 100 | 8 | (4-10) | 96 | 8 | (1-10) | 0.3454 |
| Stress intolerance | 100 | 8 | (3-10) | 96 | 5 | (1-10) | **0.0420** |
| Muscle pain | 84 | 5 | (1-10) | 83 | 4 | (1-9) | 0.6906 |
| Headache | 95 | 5 | (1-9) | 87 | 5 | (1-10) | 0.4526 |
| Joint pain | 89 | 3 | (1-10) | 78 | 3 | (1-9) | 0.4640 |
| Memory/word finding problems | 100 | 5 | (2-7) | 70 | 5 | (1-8) | 0.6422 |
| Concentration impairment | 100 | 6 | (3-9) | 91 | 5 | (1-9) | 0.2879 |
| Mental fatigue | 100 | 6 | (4-10) | 100 | 7 | (2-10) | 0.6258 |
| Visual disturbances | 63 | 3 | (1-6) | 48 | 1 | (1-6) | 0.4373 |
| Palpitations | 89 | 5 | (1-10) | 70 | 4 | (1-9) | 0.4448 |
| Dizziness when standing up | 84 | 5 | (1-10) | 83 | 4 | (1-8) | 0.5256 |
| Dizziness when walking | 68 | 3 | (1-7) | 61 | 2 | (1-9) | 0.3169 |
| Sleep disturbances | 89 | 7 | (1-10) | 83 | 6 | (1-10) | 0.9999 |
| Temperature sensitivity | 79 | 5 | (1-8) | 48 | 1 | (1-8) | **0.0241** |
| Sensitivity to light | 84 | 5 | (1-10) | 52 | 2 | (1-7) | **0.0143** |
| Sensitivity to noise | 89 | 5 | (1-10) | 70 | 3 | (1-9) | **0.0290** |
| Breathing disorders | 79 | 5 | (1-10) | 70 | 5 | (1-10) | 0.6710 |
| Irritable bowel | 79 | 5 | (1-9) | 48 | 1 | (1-10) | 0.1821 |
| Fever | 21 | 1 | (1-10) | 17 | 1 | (1-3) | 0.8714 |
| Painful lymph nodes | 32 | 1 | (1-9) | 30 | 1 | (1-7) | 0.9186 |
| Sore throat | 63 | 3 | (1-7) | 57 | 2 | (1-7) | 0.4096 |
| Flu-like feeling | 79 | 5 | (1-8) | 70 | 3 | (1-10) | 0.0944 |

CCS/CFS = chronic COVID syndrome/chronic fatigue syndrome; CCS = chronic COVID syndrome without fulfilling CFS criteria; PEM = post- exertional malaise.

Patients with CCS/CFS reported significantly more stress intolerance (p=0.042) and hypersensitivity to temperature (p=0.024), noise (p=0.029) and light (p=0.014) (**Fig. 3 and Table 2**). All other symptoms were not significantly different in frequency and severity between CCS/CFS and CCS.

**Fig. 3.**
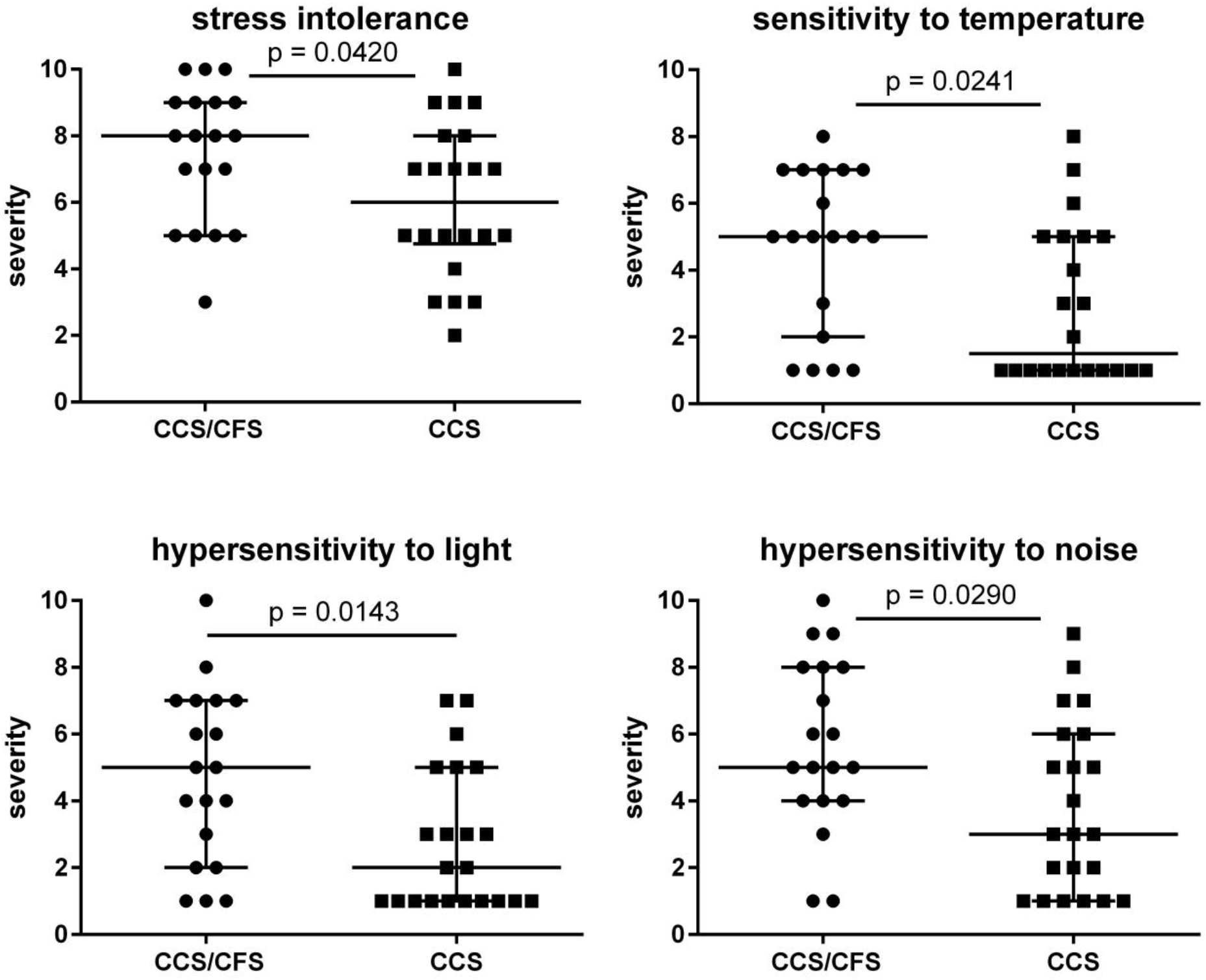
Severity of symptoms. Severity of symptoms was assessed on a 1 – 10 Likert scale (none to severest). Median, range and single values are shown in 19 CCS/CFS and 23 CCS patients.

### Autonomic dysfunction

The majority of patients suffer from autonomic dysfunction assessed by COMPASS-31 score with moderate (20–40) symptoms in 21 and severe (40–60) in 11 patients. Severity of symptoms was not significantly different between CCS/CFS and CCS. The COMPASS total score and subdomains of orthostatic, gastrointestinal, vasomotor, pupillomotor, secretory and bladder symptoms are listed in **Table 3**.

**Table 3:** COMPASS total score and subdomains.

| COMPASS 31 | max. value | CCS/CFS |  |  | CCS |  |  |
| --- | --- | --- | --- | --- | --- | --- | --- |
|  |  | % | median | range | % | median | range |
| **total** | 100 | 100 | **39.37** | (7.1-62.2) | 100 | **26.79** | (2.5-54.0) |
| **orthostasis** | 40 | 84 | **24.00** | (0-40) | 76 | **16.00** | (0-40) |
| **vasomotor** | 5 | 26 | **0.00** | (0-4.2) | 10 | **0.00** | (0-4.2) |
| **secretomotor** | 15 | 63 | **4.29** | (0-12.9) | 57 | **2.14** | (0-10.7) |
| **gastrointestinal** | 25 | 89 | **6.25** | (0-15.2) | 95 | **6.25** | (0-16.1) |
| **bladder** | 10 | 47 | **0.00** | (0-3.3) | 33 | **0.00** | (0-2.2) |
| **pupillomotor** | 5 | 95 | **1.67** | (0-3.3) | 86 | **1.33** | (0-3.0) |

CCS/CFS = chronic COVID syndrome/chronic fatigue syndrome; CCS = chronic COVID syndrome without fulfilling CFS criteria; COMPASS 31 = Composite Autonomic Symptom Score

### Hand grip strength (HGS)

Muscle fatigue and fatigability were assessed by ten maximum HGS, which were repeated after 60 minutes (Fmax and Fmean1/2). Compared to reference values for age-matched healthy females most patients were below the cut-off values for Fmax1/2 and Fmean1/2 discriminating HC from ME/CFS (own manuscript in revision). Patients with CCS/CFS had significantly lower Fmax2 and Fmean1/2 compared to CCS (**Fig. 4**). Values for male patients are not shown due to low numbers.

**Fig. 4.**
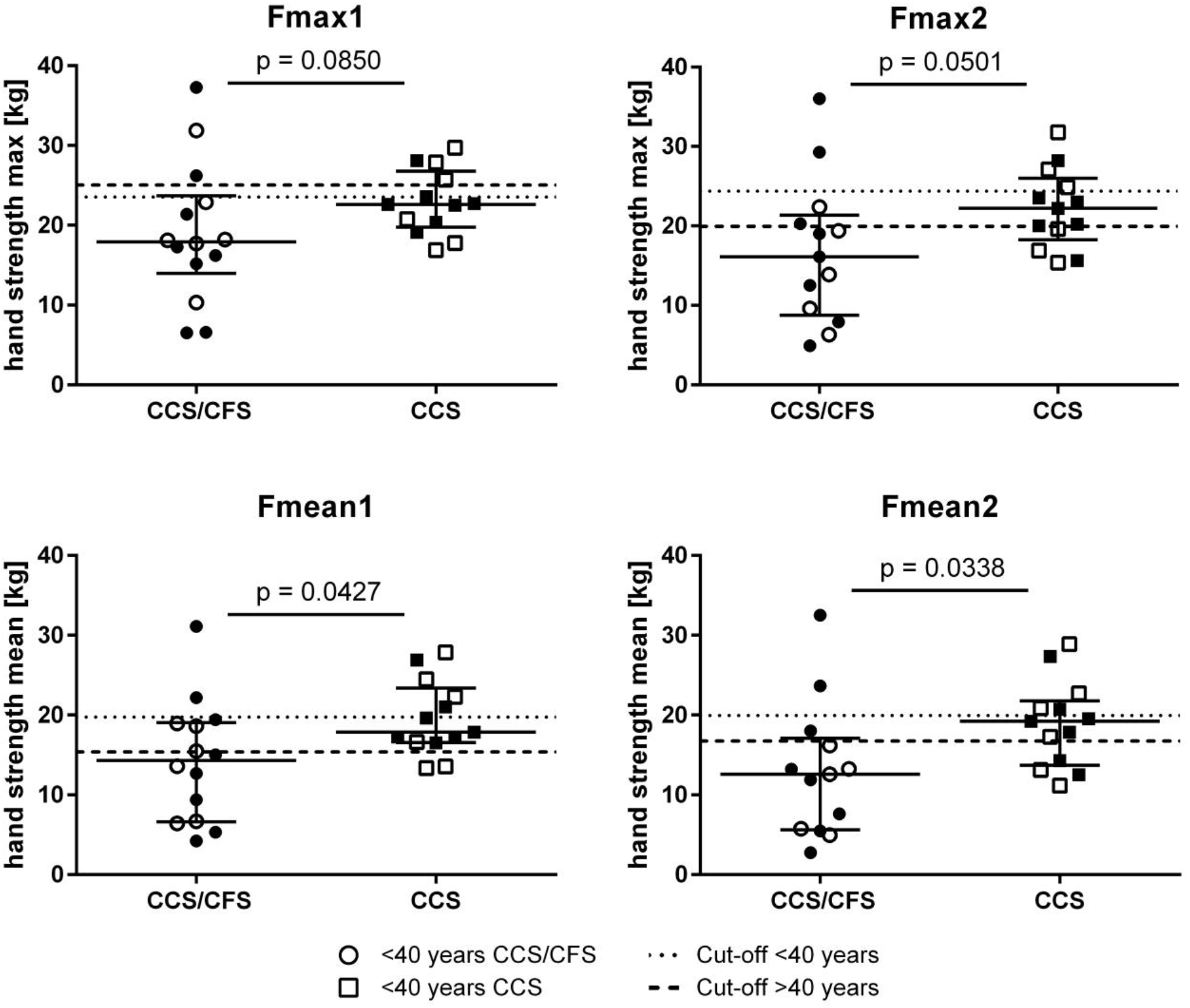
Hand grip strength (HGS) Fmax1 and fmean1 (of 10 repeat Fmax) and repeat assessment after 60 minutes (Fmax2 and Fmean2). Median. range and single values are shown in 14 female CCS/CFS and 15 female CCS patients. Cut-off values of AUC reference values for age-matched healthy females are shown as dashed lines (<40 years black dots and narrower dashed lines. >40 years white dots and wider dashed lines.).

### Sitting and standing heart rate and blood pressure

Heart rate and systolic and diastolic blood pressure sitting and after five minutes standing in female patients is shown in **Fig.5**. Patients with CCS/CFS had a significantly lower increase in systolic and diastolic blood pressure at standing compared to CCS. Seven patients with CFS and six with CCS had a blood pressure >140/90 sitting. Four patients with CCS/CFS and no patient with CCS fulfilled diagnostic criteria for postural tachycardia syndrome (POTS).

**Fig 5.**
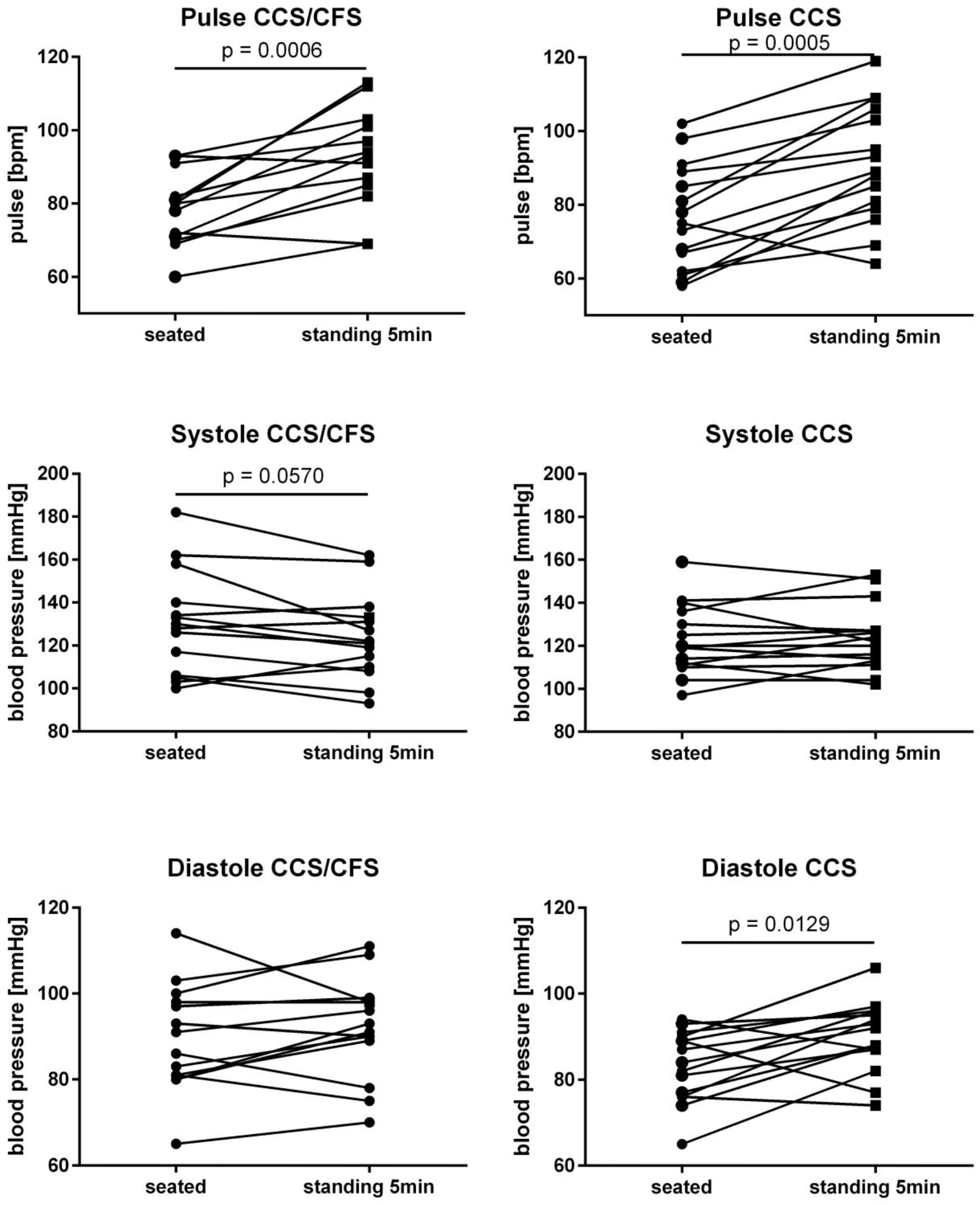
Sitting and standing heart rate and blood pressure. Heart rate and systolic and diastolic blood pressure sitting and after five minutes standing. Single values are shown in 14 female CCS/CFS and 15 female CCS patients.

### Laboratory parameters

Supplementary table S2 lists the laboratory values, which were out of normal range in a subset of patients. Mild lymphopenia with diminished CD4 (range from 0.31-0.49/nl. n=5) and or CD8 cells (n=11) was found rather frequently. Further MBL deficiency was more frequent with 22% in all patients compared to 5% in the general population.^20^ While CrP was slightly elevated with 7.1 and 7.6 mg/l in two patients only, IL-8 in erythrocytes indicating elevated levels during the last three to four months was above the normal value in 43% of the patients. IgE was elevated in 21% of patients. Elevated ANA of 1:160 - 1:1280 was found in three patients in the CCS/CFS and in six patients in the CCS cohort. The patients were seen by a rheumatologist with no evidence for a rheumatic disease and ENA antibodies were not detectable in eight of eight patients. Deficiencies of Vitamin D and folic acid were found in 17% and 14% of patients. NT-pro BNP was only slightly elevated in three patients (103-111ng/l). There were no significant differences comparing laboratory findings in CCS/CFS vs CCS.

## DISCUSSION

In this study we provide evidence that COVID-19 can trigger a severe chronic syndrome with the hallmark of fatigue and exertion intolerance. Half of the patients reported here fulfill the CCC for ME/CFS.^18^ The others did not fulfill the CCC mostly due to shorter duration of PEM lasting for two to ten hours only.^8^ Due to the overlap with ME/CFS we suggest Chronic COVID-19 Syndrome as an appropriate terminology in accordance with another report.^21^ In our study, patients fulfilling the diagnostic criteria for ME/CFS had more severe fatigue and functional disability and reported more severe stress intolerance and hypersensitivity. Moreover, hand grip strength was lower in this subgroup referred as CCS/CFS, but also considerably impaired in many of the non-CFS patients referred as CCS. Several diagnostic criteria have been proposed for use in ME/CFS, of which CCC are recommended for diagnosis confirmation in secondary care and in research.^22^ Here the severity and duration of PEM is a key diagnostic criterion. In contrast to the original minimum length of 24 hours of PEM required by the CCC we set the duration criterion at 14 hours, which was shown to yield the highest diagnostic sensitivity and specificity to discriminate patients with ME/CFS from patients with fatigue due to other chronic illness.^8 12^ There are simpler criteria including the IOM and the CDC-1994/Fukuda criteria but they should be used for screening purposes only as both lack key symptoms required in CCC for diagnosis.^23 24^ Of note, use of such criteria would have classified more patients from our study as ME/CFS.

ME/CFS is a debilitating disease leading to vast social, economic and individual impairments.^22^ People with ME/CFS have been struggling for decades to be recognized as having a serious and debilitating illness as many physicians are not familiar to diagnose and treat this disease. Despite the less severe expression of some symptoms in the CCS subgroup, most of the patients are severely impaired in daily life, too. Based on average Bell score about two thirds of patients require a reduced work schedule or are unable to work. This finding is in accordance with the recent report of a patient survey from long COVID at seven months.^3^

Health sequelae of long COVID-19 may be multiple. The most relevant are post intensive care syndrome, pulmonary impairment, neurological deficits and posttraumatic stress disorder. In our patient cohort of younger patients with mostly mild COVID-19, we have, however, no evidence for potential confounding organ impairment or major depressive or anxiety diseases in accordance with other reports.^25 26^ A study from a pulmonary center recently described that patients with normal lung function three months after recovery from acute mild COVID-19 and with normal lung function exhibited more fatigue and impairment of physical functioning and as well as quality of life than patients who had moderate-to-critical COVID-19.^26^ Furthermore, in this study a minority of patients had evidence for depression or anxiety in line with our data, providing evidence that despite a high illness burden mental health is not relevantly impaired in most patients with CCS.

There is still no specific treatment for ME/CFS and the knowledge of pathomechanisms is fragmented due to little interest and research support.^27^ There is evidence of immune, autonomic and metabolic dysregulation in post-infectious ME/CFS.^13^ In line with these data, most patients in our study presented with symptoms of autonomic dysfunction. COVID-19 results in a strong inflammatory response and there is evidence for autoimmunity triggered by COVID-19.^28^ We have no evidence for ongoing overt inflammation as only two of the patients had mildly elevated CrP. Almost half of the patients had, however, elevated IL-8 levels in erythrocytes. IL-8 was significantly increased in critical compared to non-critical acute COVID-19.^29^ Elevated ANA antibodies in nine patients and the preponderance of females may indicate an autoimmune mechanism similar to ME/CFS triggered by other infections.^13-15^ MBL deficiency which has been implicated in susceptibility and course of viral infections was found more frequently in CCS/CFS in accordance with findings from a past study in ME/CFS.^20^

### Limitations of this study

Our study has several limitations. Firstly, recruitment bias may have contributed to the severity of symptoms, thus this cohort is probably not representative for all COVID-19 patients with persistent fatigue. Secondly, it is unclear, if the distinction into subgroups based on the criterion of the length of PEM indicates differences in mechanisms or merely reflect the variance of the disease spectrum. Thirdly, the low number of patients precluded detailed comparisons of phenotypes with adequate statistical power.

## CONCLUSION AND POLICY IMPLICATION

Our study provides evidence that patients following mild COVID-19 develop a chronic syndrome fulfilling diagnostic criteria of ME/CFS in a subset. We must anticipate that this pandemic has the potential to dramatically increase numbers of ME/CFS patients. At the same time it offers the unique chance to identify ME/CFS patients in a very early stage of disease, so that interventions such as pacing and coping can be applied early with a better therapeutic prognosis. Further, it is an unprecedented opportunity to understand the pathomechanisms and characterize targets for specific treatment approaches.

## Data Availability

Datset ist available upon request.

## Contributors

CSc, CK and FP developed the concept of the study. HDV gave important input into study concept and objectives HF and CK were responsible for data curation and analyses of data. CK. CSc, KW, FS, JBS, LMA, FP, BH, and CSk were involved in clinical investigation. MH and RG were involved in data transfer and patient care. TZ was involved in ethical affairs and data management. HF, CK, and CSc validated the data. HF was involved in data visualization. CSc wrote the original draft of the manuscript. CK and HF reviewed and edited the manuscript. All authors revised and approved the manuscript. The corresponding author attests that all listed authors meet authorship criteria and that no others meeting the criteria have been omitted. CSc is the guarantor.

## Funding

This work is supported by a grant from the Weidenhammer Zöbele Foundation.

## Competing interests

All authors declare that they have no conflict of interests. All authors have completed the ICMJE uniform disclosure form.

## Ethical approval

Ethical approval was given by the Ethics Committee of Charité Universitätsmedizin Berlin in accordance with the 1964 Declaration of Helsinki and its later amendments (EA2/066/20).

## Informed consent

All participants provided written informed consent.

## Transparency

The lead author (the manuscript’s guarantor) affirms that the manuscript is an honest, accurate, and transparent; that no important aspects of the study have been omitted; and that any discrepancies from the study as planned have been explained. Dissemination to participants and related patient and public communities is encouraged by open access publication and citing the study on our site https://cfc.charite.de/. We are engaging with print and internet press, television, radio, news, and documentary program makers.

## Acknowledgement

We thank Silvia Thiel for patient care and data management. We thank all patients who gave us their consent to publish their data in this st

## Supplemental material

**Supplementary table S1:**
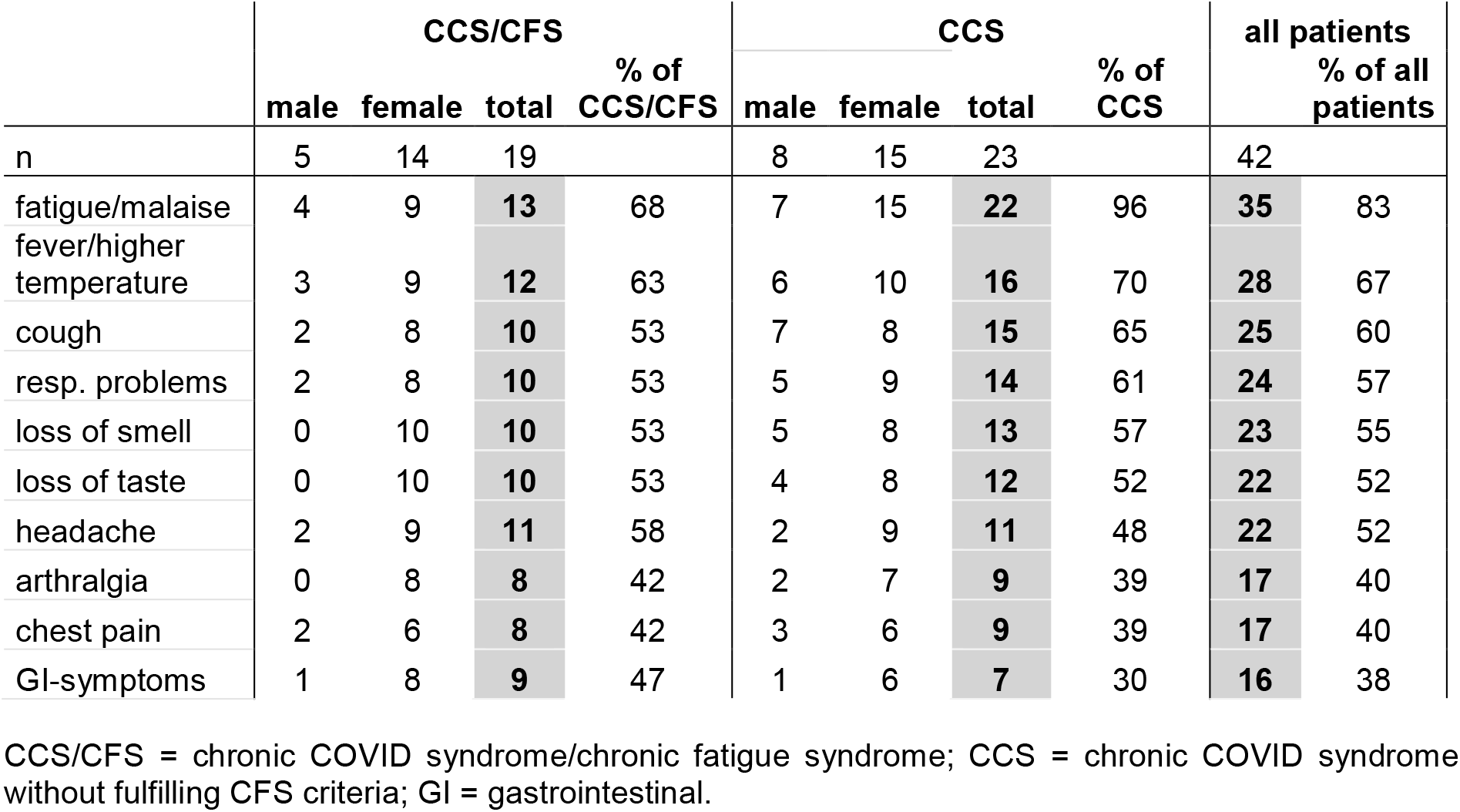
ten most frequent initial symptoms of COVID-19 reported by patients

**Supplementary table S2:** Laboratory parameters

| Lab Value | CCS/CFS (n = 19) |  | CCS (n = 23) |  |
| --- | --- | --- | --- | --- |
|  | n | % | n | % |
| NK-cell high | 1 | 5 | 0 |  |
| NK-cell low | 0 |  | 1 | 4 |
| CD-4 Cell low | 1 | 5 | 4 | 17 |
| CD-4 Cell high | 2 | 11 | 1 | 4 |
| CD-8 Cell low | 3 | 16 | 8 | 35 |
| CD-8 Cell high | 2 | 11 | 0 |  |
| MBL deficiency | 4 | 21 | 5 | 22 |
| sol. IL2 receptor high | 1 | 5 | 1 | 4 |
| IL-8 after erylyse high | 6 | 32 | 12 | 52 |
| IgA high | 2 | 11 | 1 | 4 |
| IgM high | 2 | 11 | 2 | 9 |
| IgE high | 4 | 21 | 5 | 22 |
| IgG3 low | 2 | 11 | 1 | 4 |
| IgG3 high | 1 | 5 | 0 |  |
| IgG2/4 high | 1 | 5 | 0 |  |
| IgG1 high | 0 |  | 1 | 4 |
| IgG2 high | 0 |  | 1 | 4 |
| IgG4 high | 0 |  | 2 | 9 |
| elevated CRP | 1 | 5 | 1 | 4 |
| elevated NT-pro BNP | 2 | 11 | 1 | 4 |
| low C3 | 1 | 5 | 3 | 13 |
| Vit D low | 2 | 11 | 5 | 22 |
| folic acid low | 2 | 11 | 4 | 17 |
| ANA positive (1:160) | 2 | 11 | 3 | 13 |
| ANA positive (1:320) | 0 |  | 2 | 9 |
| ANA positive (1:1280) | 1 | 5 | 0 |  |

## Notes

### Funding Statement

This work is supported by grants from the Weidenhammer Zoebele Foundation

### Author Declarations

All patients signed informed consent before study assesments. This study is part of the Pa-COVID19-Project of Charite and was approved by the Ethics Committee of Charite (Universitaetsmedizin Berlin in accordance with the 1964 Declaration of Helsinki and its later Amendments).

